# The Impact of Neurofeedback on Effective Connectivity Networks in Chronic Stroke Patients

**DOI:** 10.1101/2020.05.04.20087163

**Authors:** Lioi Giulia, Veliz Adolfo, Coloigner Julie, Duché Quentin, Butet Simon, Mathis Fleury, Emilie Leveque-Le Bars, Elise Bannier, Anatole Lécuyer, Christian Barillot, Isabelle Bonan

## Abstract

Stroke is a complex motor disease that not only affects perilesional areas but also global brain networks in both hemispheres. Neurofeedback (NF) is a promising technique to enhance neural plasticity and support functional improvement after stroke by means of brain self-regulation. Most of the studies using NF or brain computer interfaces for stroke rehabilitation have assessed treatment effects focusing on motor outcomes and successful activation of targeted cortical regions. However, given the crucial role of large-scale networks reorganization for stroke recovery, it is now believed that assessment of brain connectivity is central to predict treatment response and to individualize rehabilitation therapies. In this study, we assessed the impact of EEG-fMRI NF training on connectivity strength and direction using a Dynamic Causal Modeling approach. We considered a motor network including both ipsilesional and contralesional premotor, supplementary and primary motor areas. Our results in nine chronic stroke patients indicate that NF upregulation of targeted areas (ipsilesional SMA and M1) not only modulated activation patterns, but also had a more widespread impact on fMRI bilateral motor networks. In particular, inter-hemispheric connectivity between premotor and primary motor regions decreased, and ipsilesional self-inhibitory connections were reduced in strength, indicating an increase in activation during the NF motor task. To the best of our knowledge, this is the first work that investigates fMRI connectivity changes elicited by training of localized motor targets in stroke. Our results open new perspectives in the understanding of large-scale effects of NF training and the design of more effective NF strategies, based on the pathophysiology underlying stroke-induced deficits.

## Introduction

A growing body of evidence suggest that post-stroke motor deficits are related to altered interactions between brain areas also remote from the stroke lesion [1]. Indeed, localized structural lesions resulting from stroke are likely to affect brain connectivity through the brain. As a consequence, recovery from stroke depends on structural and functional networks reorganization [2]–[5]. These findings are in line with the increasingly influential thesis that most of complex biological diseases, such as motor disorders [1], psychiatric disorders [6] or Alzheimer disease [7] are related to the dysfunction of complex brain networks [8].

In stroke in particular, several studies have used brain functional neuroimaging techniques such as positron emission tomography (PET), functional Magnetic Resonance imaging (fMRI) and electroencephalography (EEG) to investigate changes in brain activity [9], [10]. More recently, connectivity-based analyses have shed new light into the pathophysiology underlying stroke-induced deficits [4], [11], [12]. A general trend observed in these studies is that stroke induces changes in both hemispheres and causes a disruption of ipsilesional connectivity [13]. Some of these connectivity-based approaches have gone further and investigated the effect of interventions (such as physical or robotic assisted therapy) on cortical network reorganization, and its link to motor function recovery [14]. However, the impact of specific therapies on motor connectivity in stroke remains to be investigated. Assessment of changes in connectivity networks induced by treatment is now considered to be a promising way to individualize therapies and enhance rehabilitation outcome after stroke [1].

There are different ways of estimating brain connectivity from neuroimaging data. Structural connectivity refers to anatomical connections between brain regions and is most commonly estimated using Diffusion Tensor Imaging (DTI). Functional connectivity (FC) is defined as the statistical dependence among measurements of neural activity [15] and it is usually inferred through correlations among neurophysiological signals. Effective connectivity (EC) estimates the influence that one neuronal system exerts on another. EC is intrinsically directed, but does not necessarily implies a direct coupling mediated by anatomical connections. Connectivity studies in stroke have relied both on FC [16], [17] and EC [3], [18], [19]. Among EC estimators, those based on or Dynamic Causal Modeling (DCM) are particularly robust for fMRI data analysis, while lag-based approach (i.e. Granger causality) may perform poorly if the hemodynamic effects are not properly taken in account [20]. More specifically, DCM modeling for fMRI includes a hemodynamic model that is deconvoluted in the estimation procedure (i.e. model inversion) to obtain a direct measure of neuronal networks causality [21].

Among the set of therapies proposed for stroke rehabilitation, Neurofeedback (NF) is gaining increasing attention since it potentially promotes plasticity of perilesional areas by means of brain self-regulation. Recent works have investigated the potential of NF (or Brain Computer Interfaces, BCI) for motor rehabilitation after stroke as an alternative or in addition to traditional physical therapies [22]–[25]. In these studies, patients perform motor imagery (MI) of the affected limb, which is a promising mental therapy to restore motor activation. Even if there is no clinical evidence of the benefit of NF over MI for stroke rehabilitation, NF was shown to enhance the efficacy of MI training, by eliciting more specific brain patterns [26], [27]. In some applications, the NF paradigm allows to integrate the feedback in an orthosis to support the movement of the affected limb, thus closing the sensorimotor loop [28], [29].

The majority of these NF approaches have relied on one imaging technique, mainly electroencephalographic (EEG) recordings. EEG-NF applications are generally based on the training of sensorimotor rhythms in motor regions ipsilateral to the stroke lesion [30]-[32]. Similarly, more recent fMRI-NF studies have promoted upregulation of ipsilesional motor areas or motor system connectivity [24], [33]–[35]. In a study involving 10 chronic stroke patients, an enhancement of the FC (node degree) of preserved ipsilesional motor areas with NF led to a significant increase in motor function. This was not the case in a control condition where patients enhanced FC of a brain area not directly implicated in motor function [34].

Building on the pioneering work of Zotev and collaborators [36], [37], our group was the first to integrate EEG and fMRI for bimodal NF for motor training, with the rationale of providing a more specific feedback, combining high temporal (EEG) and spatial (fMRI) resolution [38]. We have also shown the feasibility of EEG-fMRI NF and its potential to promote upregulation of ipsilesional motor regions in a pilot study involving chronic stroke patients [25]. In this previous work, we have investigated NF training effects focusing on the activation of localized target cortical areas. In the present study, we characterize global changes elicited by NF training on EC networks in chronic stroke patients undergoing a longer NF training protocol. We assessed fMRI connectivity changes using a DCM approach and considered a motor network including bilateral premotor, supplementary and primary motor areas (PMC, SMA, M1). We believe that our results bring new insight into large-scale effects of NF training in stroke and on the potential of NF in driving maladaptive networks reorganization.

## Methods

### Participants

Nine chronic stroke patients (mean age 59 years, age range: 37-77 years, 3 females) were included in the study. Patients had mild to severe hemiparesis of the upper limb (Upper extremity Fugl-Meyer score in the range 26-55) with variable lesion characteristics (Table 1) but were clinically stable, at more than one year from the stroke episode (33.2 ± 16.5 months). Integrity of the corticospinal tract (CST) was an inclusion criterion. Patients for whom the CST fractional anisotropy asymmetry index exceeded the 0.15 threshold [39] were not enrolled. More details about diffusion imaging processing and CST segmentation are given in Supplementary Material. All participants gave their written informed consent and the study was approved by the institutional review board and registered (NCT03766113). The registered study is a simple-blind randomized controlled trial including a control group undergoing MI training without NF. For the purpose of this ancillary work focusing on NF training effects on effective connectivity, only patients in the NF group were considered.

**Table 1.**
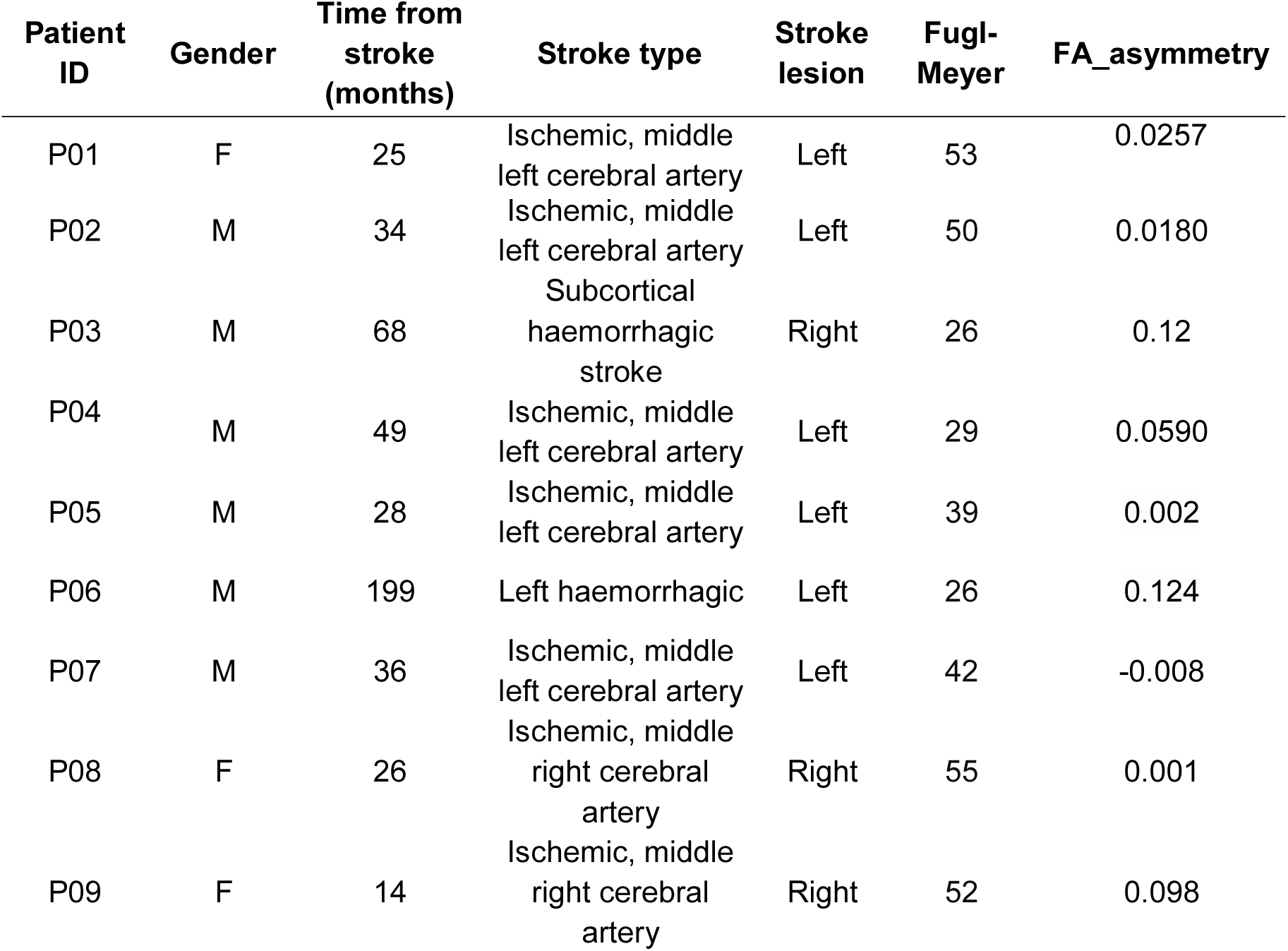
Patients demographic and stroke characteristics.

| Patient ID | Gender | Time from stroke (months) | Stroke type | Stroke lesion | Fugl-Meyer | FA_asymmetry |
| --- | --- | --- | --- | --- | --- | --- |
| P01 | F | 25 | Ischemic, middle left cerebral artery | Left | 53 | 0.0257 |
| P02 | M | 34 | Ischemic, middle<br>left cerebral artery | Left | 50 | 0.0180 |
| P03 | M | 68 | Subcortical<br>haemorrhagic<br>stroke | Right | 26 | 0.12 |
| P04 | M | 49 | Ischemic, middle<br>left cerebral artery | Left | 29 | 0.0590 |
| P05 | M | 28 | Ischemic, middle<br>left cerebral artery | Left | 39 | 0.002 |
| P06 | M | 199 | Left haemorrhagic | Left | 26 | 0.124 |
| P07 | M | 36 | Ischemic, middle<br>left cerebral artery | Left | 42 | -0.008 |
| P08 | F | 26 | Ischemic, middle<br>right cerebral<br>artery | Right | 55 | 0.001 |
| P09 | F | 14 | Ischemic, middle<br>right cerebral<br>artery | Right | 52 | 0.098 |

### Experimental Protocol

Participants underwent a NF protocol alternating bimodal EEG-fMRI NF (N=5) and unimodal EEG-NF sessions (N=9) over a period of five weeks (Figure 1). Bimodal NF sessions included a calibration and three NF training blocks alternating epochs of rest (20 s) and NF-MI training (20 s) (Figure 1). Similarly, the unimodal EEG NF sessions consisted of a calibration period followed by three NF runs with a block-design alternating rest and task during 5 min, with an amount of training time and protocol structure equivalent to the bimodal training sessions. Patients were informed at inclusion, verbally and by an explanatory note, about the goals and the timeline of the study. Instructions were repeated before each training session and oriented the patients towards a kinesthetic MI of the affected upper limb, without mentioning a specific strategy.

**Figure 1.**
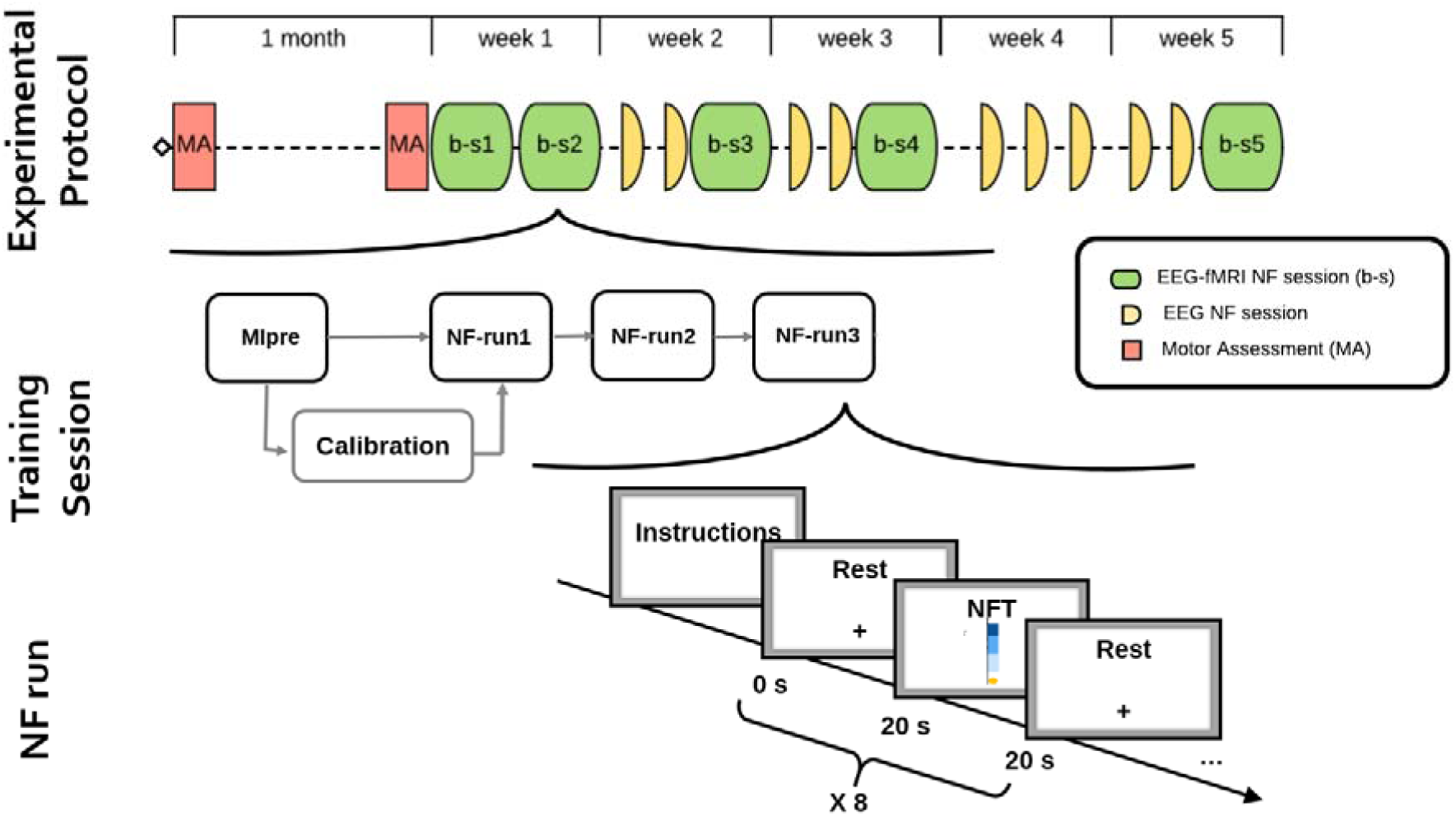
A schematic of the experimental protocol. Motor assessment (MA) and NF training sessions timeline is shown in the first row (alternation of bimodal EEG-FMRI b-s* and unimodal EEG NF session). The second row is a schematic of the typical training session with a first Motor Imagery run for calibration purpose and 3 NF training runs. Finally, the third row shows the block design alternating 20 s rest and 20 s NF training of a NF run.

Imaging was performed using a Siemens 3T Prisma scanner running VE11C and a 64-channel MR-compatible EEG system from Brain Products (Brain Products GmbH, Gilching, Germany). Functional MRI data were acquired with echo-planar imaging (EPI) with the following parameters: repetition time (TR)/echo time (TE) = 1,000/23 ms, FOV = 230 × 230 mm2, 16 4-mm slices, voxel size = 2.2 × 2.2 × 4 mm3, matrix size = 105 × 105, flip angle = 90°. For each session, a high-resolution 3D T1 MPRAGE sequence was acquired with the following parameters: TR/TI/TE = 1,900/900/2.26 ms, parallel imaging with GRAPPA 2, FOV = 256 × 256 mm2 and 176 slabs, voxel size = 1 × 1 × 1 mm3, flip angle = 9°. In order to assess the asymmetry between the ipsilesional and controlesional CST, diffusion imaging (TR/TE=11000/99ms, FOV 256×256mm^2^, 60 slices, matrix 128×128, voxel size, 2×2×2 mm^3^, 30 directions, b=1000s/mm^2^) was performed before inclusion. During rest, a cross was displayed on the screen and participants were asked to concentrate on the cross and rest. During the task, a visual feedback in form of a ball moving in a one-dimensional gauge proportionally to the average of the EEG and the fMRI features was presented. Unimodal EEG-NF sessions were performed using the Mensia Modulo (MENSIA TECHNOLOGIES) hardware solution, equipped with an 8-channel EEG cap. Patients were exposed to the same feedback metaphor on a computer screen. More details about the NF platform performing real-time EEG-fMRI processing, the unimodal NF sessions and the experimental protocol are given in [25], [38], [40] and in the attached Supplementary Material.

### Calibration and online NF calculation

At the beginning of each training session, a MI task without NF was performed to calibrate the fMRI signal. Functional MRI data were pre-processed for motion correction and slice-time correction and realigned with the structural scan. After spatial smoothing with a 6mm FWHM Gaussian kernel, a general linear model (GLM) analysis was performed. The calibration activation map was used to define two regions of interest (ROIs) to calculate NF scores for the subsequent NF task. Boxes of 9×9×3 voxels (20×20×12 mm^3^) centered around the peak of activation in the ipsilesional SMA and M1 were considered. As detailed in a previous work [25], we proposed an adaptive rewarding strategy that more importantly weighted SMA at the beginning of the training and then guided the patients towards activation of the ipsilesional M1. The fMRI NF score was therefore calculated as a weighted combination of the activity in the SMA and M1 ROIs, with weights changing at each training session. In particular the fMRI-NF score was calculated as the difference between percent signal change in the two ROIs (SMA and M1) and a background slice whose activity is not correlated with the NF task, in order to reduce the impact of global signal changes [41].

For EEG calibration, 18 EEG channels located over the motor regions were considered to estimate a Common Spatial Pattern (CSP) filter [42]. For online EEG-NF computation, the band power (BP) in the 8-30 Hz band was computed from filtered data and normalized with respect to the power of the previous rest block (event related desynchronization). For the sake of brevity of this manuscript focusing on fMRI effective connectivity analysis, we have provided more details about calibration and NF calculation procedure in Supplementary Material. A CRED-nf checklist with details about experimental design [43] is also available as Supplementary File 1 together with a table summarizing real-time signal processing steps, according to the COBIDAS-inspired template [44] (Table S1, Supplementary Material).

### fMRI preprocessing

In order to assess the effect of the multi-session NF training protocol on EC patterns, we considered fMRI time-series of the first (b-s1) and last (b-s5) bimodal NF training sessions. BOLD series were pre-processed and processed with Matlab and SPM12 (Wellcome Department of Imaging Neuroscience, UCL, London, UK). A slice timing correction was applied to correct for timing differences within volumes. Functional volumes were then registered to the mean volume to compensate for subject motion within the series and ensure voxel-to-voxel correspondence across time. The mean fMRI image was co-registered with the subject structural image. The anatomical volume was in turn segmented and nonlinearly transformed to the reference Montreal Neurological Institute (MNI) space. The estimated normalization deformations fields were later applied to functional data that were finally smoothed using a 3D Gaussian kernel of 6mm FWHM.

An offline data quality control was performed to assess for the impact of head movement artifacts on data quality and NF performances. Motion parameters were estimated and used to perform a post-hoc correlation analysis with the NF task. To this end and for each bimodal session a Framewise Displacement (FD) was computed from the six realignment parameters as proposed by Power et al. [45]. FD outliers were identified using the spmup tool within the QAP Package [46]. Pearson correlation analysis between the FD time-series and the NF task was assessed.

At the subject level, each session was modeled using a three-run general linear model (GLM), where each NF block was modeled by convoluting a 20s boxcar function with the standard hemodynamic response function (HRF) to build a NF task regressor. The estimated motion parameters and a run mean were also entered in the design matrix as covariates of no interest. The average brain response across the three runs during the NF task was calculated to get individual contrast maps. The individual contrast maps of the three patients with right lesions were flipped to create an artificial group of 9 patients with left hemispheric lesions. These maps were entered into a training session specific second level analysis GLM to evaluate group activation maps at each NF training session, allowing to monitor the effect of NF on brain activation.

### ROI definition and time series extraction

In order to discard task independent noise, representative time series for the selected ROIs were extracted by selecting the voxels which, in the first level GLM analysis, exceeded the statistical threshold for NF contrast (p<0.01), and were located within an apriori mask for bilateral SMA, PMC and M1. These masks were defined using the Human Motor Area Template (HMAT) atlas [47]. The SMA mask included both preSMA and SMA HMAT ROIs, and the PMC included ventral and dorsal PMC. To adapt the selected ROIs to individual responses, we extracted the first principal component of the time series from all voxels within 8-mm spheres around local activation maxima [48]. As for whole brain analysis, ROIs time series were flipped in the case of patients with a right hemisphere lesion (N=3) for the sake of the group analysis (in the group results of the affected hemisphere is thus presented on the left).

### Dynamical causal modeling (DCM)

DCM is a hypothesis-based technique that describes how observed fMRI responses are generated using a set of differential equations and represents one of the most common framework for the analysis of fMRI effective connectivity [49]. These differential equations describe how experimental stimulation (input) produces changes in neural activity and induces changes in the output (i.e. the observed fMRI data) through an hemodynamic model [50]. DCM models include parameters, such as the strength of coupling between the ROIs, i.e. effective connectivity, which are estimated from the data using a variational Laplace approach [51].

In the current DCM study, we proposed a motor network model consisting of six regions with bidirectional endogenous connections among them all (PMC_L_, PMC_R_, SMA_L_, SMA_R_, M1_L_, and M1_R_). These regions were selected on the basis of the evidence of their role in MI and previous connectivity stroke studies [4], [14], [52]. In this study we focused on endogenous connectivity analysis (DCM matrix A) [48] and assessed how its strength is modulated by the NF training.

In the first step of the analysis, a Bayesian model selection procedure for each subject was performed to estimate the model that best matched the measured fMRI data. The basic 6 ROIs model was elaborated into 10 more different models depending upon which premotor region (SMA and PMC) was modulated by the external experimental input (NF task) and a ‘null’ model with no modulation. The different models (represented in Figure S2 in Supplementary Material) were compared and an optimal DCM model for each subject and session was identified using the Bayesian model selection.

After all DCM models were estimated and every subject connectivity strengths inferred, we performed a group level analysis to assess differences and commonalities between patients using the recently introduced Parametrical Empirical Bayes (PEB) approach [53]. PEB assumes that all the subjects have the same network architecture, differing only in the strength of the connections. To test hypotheses about between-subjects effects, individual differences in coupling parameters are decomposed into hypothesized group level effects, called regressors or covariates, and unexplained variance or random effects. Since the aim of this study was to assess the effect of the NF training on the EC strength, we introduce this covariate of interest in the PEB analysis. In order to exclude other factors independent from the training, the Fugl-Meyer score, FA asymmetry index, type of stroke and time from stroke were also included as regressors in the between-subjects analysis. We applied PEB to each model and extracted the free energy, which is an approximation of the log-evidence of the model that considers both accuracy and complexity [51], compared them and selected a winning model. The winning model was then considered for the posterior analysis and the group level results.

Finally, after having identified the winning model, an automatic search over reduced models was performed by means of Bayesian Model Reduction (BMR) [53]. To summarize the results over all the models, a Bayesian Model Average (BMA) was computed [54] by averaging the parameters from different reduced models, weighted by the model posterior probabilities. When showing summary BMA results, we only considered parameters with a strong evidence (i.e. posterior probability of being non-zero greater than 0.99). All the computations were performed using the DCM analysis code adapted from SPM12, described step-by-step in [48], [55] and available at https://github.com/pzeidman/dcm-peb-example. Other scripts used for fMRI and DCM analysis in this paper can be provided by the authors upon request. More details about code availability are also given in Supplementary Material.

## Results

### fMRI activation analysis

The motion artifacts analysis revealed that in 5 out of 45 training sessions the percentage of head motion outliers was higher than 10%. Also, in 11% of sessions (6 out of 45) the Pearson correlation between the head motion and the NF task regressor was higher than 0.25 (in absolute value). These findings are similar to those reported in our previous study on 30 healthy volunteers [56]. Additional details and results of the offline data quality check are given in Supplementary Material (Table S1 and Figures S1 and S2).

Figure 2 presents group activation maps (N=9) in the first (b-s1) and last (b-s5) NF training session. Reported statistical maps show activations exceeding a voxel-level uncorrected threshold of p<0.05. Average BOLD activation maps show significant activations of the premotor cortex and supplementary motor areas. The bilateral posterior parietal cortex (PPC), that plays an important role in visuo-motor coordination [57], was also recruited during the MI NF task. In general, patients robustly activated mainly the bilateral SMA during the first session (b-s1), while they showed also activation of the bilateral PMC and ipsilesional M1 in the last NF session (b-s5).

**Figure 2.**
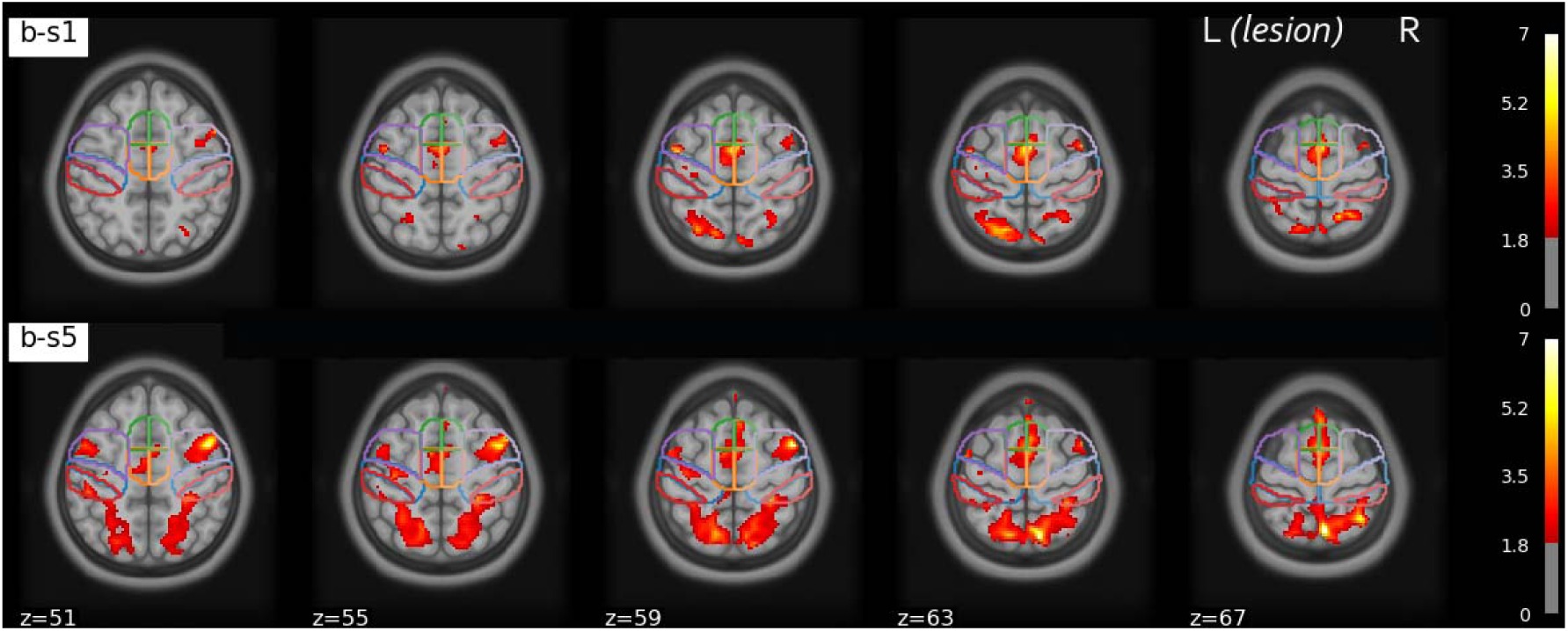
Group activation maps in the first (b-s1) and last (b-s5) training session in MNI coordinates (p<0.05, uncorrected). The outline of the motor areas of interest based on the HMAT atlas is indicated: preSMA (green) SMA (orange), PMC (purple), M1 (blue) and Sensory motor cortex (red). The lesional hemisphere is on the left (results for the patients whose lesion was in the right hemisphere were flipped for the sake of comparison).

### DCM analysis: changes in endogenous connectivity

The subject average percentage variance explained by the model is 10.5% (SD 7.3%): it compares to other results in literature [48] but is smaller. This is to be expected in the case of simultaneous EEG-fMRI imaging considered the decrease in fMRI signal to noise ratio due to the presence of EEG electrodes [58]. In most of the subjects, coupling parameters were nontrivial, with 90% credible intervals that excluded zero, indicating that there was useful information in the data pertaining to the NF experimental effects. The PEB free energy, considered to a proxy for the log model evidence, was of −1.012 × 10^5^ (a graph reporting the free energy corresponding to the 10 DCM models is reported in Supplementary Material, Figure S4).

Endogenous connectivity patterns elicited during MI NF obtained from the BMA analysis are schematically represented in Figure 3 and the corresponding values are listed in Table 2. Coupling parameters in average connectivity patterns (i.e. common to all subjects and sessions) quantify the EC from one area over another in Hz. Negative coupling rates indicate that the source region has an inhibitory effect on the activity of the target region (in red in Figure 3), while positive EC values indicate an excitatory effect (in green in Figure 3). Self-connection link scale up or down the default value of 0.5 Hz [48] therefore positive self-connections values indicate increased self-inhibition while negative values indicate activation of the specific region. As shown in Figure 3A, experimental driving inputs (NF MI task) had a positive influence on bilateral premotor areas: this is consistent with the recruitment of PMC for a MI task [59]. The average EC results indicate a fully connected motor network, with mainly excitatory coupling, involving both hemispheres. Connectivity between contralesional and ipsilesional motor areas was also elicited during the MI task of the affected limb. Self-connection values were negative for all ROIs, indicating a significant disinhibition, or equivalently an activation, of all the considered motor regions during the NF MI task.

**Table 2.** Bayesian Model Average of PEB parameters: Average endogenous connectivity and significant changes in connectivity strength as a result of the NF training.

| Source | Target | Commonalities | Effect of Treatment |
| --- | --- | --- | --- |
| SMA<sub>L</sub> | SMA<sub>L</sub> | -0.3616 | -0.1339 |
| SMA<sub>L</sub> | PMC<sub>L</sub> | -0.0000 | 0.0893 |
| SMA<sub>L</sub> | M1<sub>L</sub> | -0.0000 | 0.0000 |
| SMA<sub>L</sub> | SMA<sub>R</sub> | 0.2396 | 0.1784 |
| SMA<sub>L</sub> | PMC<sub>R</sub> | 0.0000 | 0.0856 |
| SMA<sub>L</sub> | M1<sub>R</sub> | -0.0902 | 0.1140 |
| PMC<sub>L</sub> | SMA<sub>L</sub> | 0.1077 | -0.1608 |
| PMC<sub>L</sub> | PMC<sub>L</sub> | -0.4562 | 0.0000 |
| PMC<sub>L</sub> | M1<sub>L</sub> | 0.3510 | -0.2343 |
| PMC<sub>L</sub> | SMA<sub>R</sub> | 0.0904 | -0.0808 |
| PMC<sub>L</sub> | PMC<sub>R</sub> | 0.1367 | -0.1872 |
| PMC<sub>L</sub> | M1<sub>R</sub> | 0.2282 | -0.1034 |
| M1<sub>L</sub> | SMA<sub>L</sub> | 0.0000 | 0.0000 |
| M1<sub>L</sub> | PMC<sub>L</sub> | 0.0860 | 0.0890 |
| M1<sub>L</sub> | M1<sub>L</sub> | -0.3818 | -0.1635 |
| M1<sub>L</sub> | SMA<sub>R</sub> | 0.0000 | -0.1213 |
| M1<sub>L</sub> | PMC<sub>R</sub> | 0.0000 | -0.0995 |
| M1<sub>L</sub> | M1<sub>R</sub> | 0.0000 | -0.0974 |
| SMA<sub>R</sub> | SMA<sub>L</sub> | 0.1752 | 0.0460 |
| SMA<sub>R</sub> | PMC<sub>L</sub> | 0.2006 | 0.1830 |
| SMA<sub>R</sub> | M1<sub>L</sub> | 0.0830 | 0.1711 |
| SMA<sub>R</sub> | SMA<sub>R</sub> | -0.3444 | -0.1195 |
| SMA<sub>R</sub> | PMC<sub>R</sub> | 0.2365 | 0.2620 |
| SMA<sub>R</sub> | M1<sub>R</sub> | 0.0000 | 0.1753 |
| PMC<sub>R</sub> | SMA<sub>L</sub> | 0.1227 | 0.0000 |
| PMC<sub>R</sub> | PMC<sub>L</sub> | 0.0818 | -0.1786 |
| PMC<sub>R</sub> | M1<sub>L</sub> | 0.0000 | -0.0000 |
| PMC<sub>R</sub> | SMA<sub>R</sub> | 0.2782 | 0.0876 |
| PMC<sub>R</sub> | PMC<sub>R</sub> | -0.2467 | 0.1195 |
| PMC<sub>R</sub> | M1<sub>R</sub> | 0.2232 | -0.1084 |
| M1<sub>R</sub> | SMA<sub>L</sub> | -0.0972 | -0.0000 |
| M1<sub>R</sub> | PMC<sub>L</sub> | -0.1026 | -0.1469 |
| M1<sub>R</sub> | M1<sub>L</sub> | -0.0783 | 0.0000 |
| M1<sub>R</sub> | SMA<sub>R</sub> | -0.2783 | -0.1223 |
| M1<sub>R</sub> | PMC<sub>R</sub> | -0.1689 | -0.1324 |
| M1<sub>R</sub> | M1<sub>R</sub> | -0.2707 | 0.0000 |

**Figure 3.**
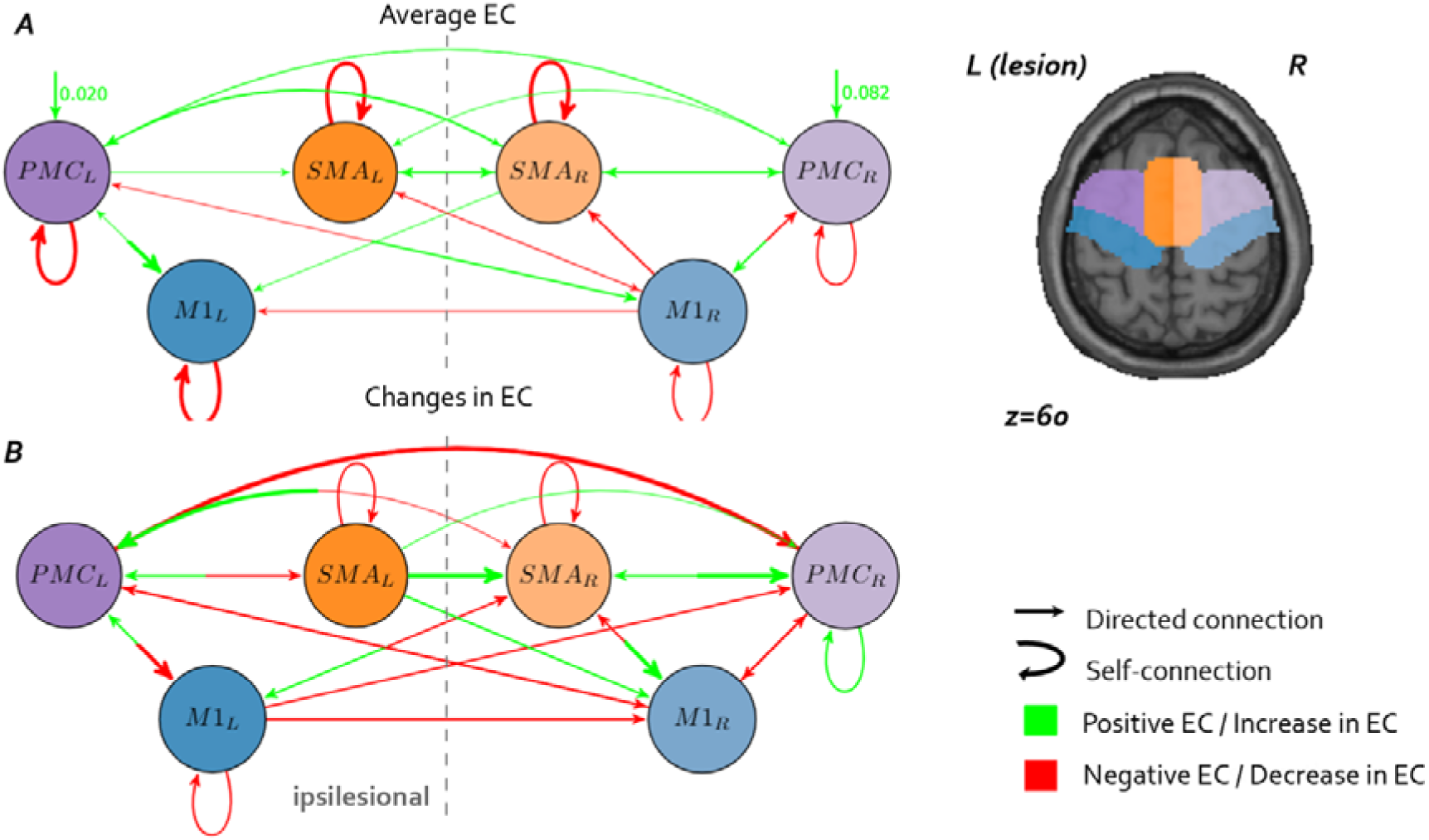
Bayesian Model Average (BMA) results. A. Connectivity network commonalities across subjects and training sessions. Arrows thickness and color code respectively for the strength and the sign of EC: green arrows represent a positive (excitatory) coupling, while red arrows indicate negative coupling. B. Significant changes (p<0.01) in EC strength after the NF training. Arrows thickness and color code respectively for the strength and the sign of EC change: green arrows represent an increase in connectivity, while red arrows indicate a decrease. The direction of links is represented by the arrow’s ending. Since EC is not symmetrical, an opposite change in connectivity can be observed between two regions. It is the case of regions connected by a “bicolor” arrow (i.e. in panel B the arrow between M1_L_ and PMC_L_ indicate an increase of the connectivity from M1_L_ to PMC_L_ and a decrease of connectivity from PMC_L_ to M1_L_).

In Figure 3B modulatory effect of the NF training on EC strength are represented, with positive connections (green) indicating an average increase in connectivity between ROIs and negative connections (red) representing a decrease in EC between the first and last NF training session. A general trend of decrease in inter-hemispheric connectivity strength can be observed, in particular between contralesional (right) and ipsilesional (left) PMC and M1. Only connections originating from SMA towards the M1 and PMC increased in strength following the NF training, while feedback connectivity from ipsilesional M1 to all other motor areas was reduced (except for the connection to ipsilesional PMC that increased). A decrease in self-inhibition strength in bilateral SMA and ipsilesional M1 was observed: this indicate a higher activation of these regions as a result of training, in line with the results of the whole brain analysis presented in Figure 2. In order to assess the variability in connectivity strength changes across subjects, we calculated the difference between individual endogenous connectivity (matrices A) in b-s1 and b-s5. Representative results for six connections (ipsilesional, contralesional and intrahemispheric EC) are shown in Figure 4 and indicate very consistent findings: individual connectivity strength have the same signs and similar amplitudes across patients. Additional results can be found in Figures S5 and S6.

**Figure 4.**
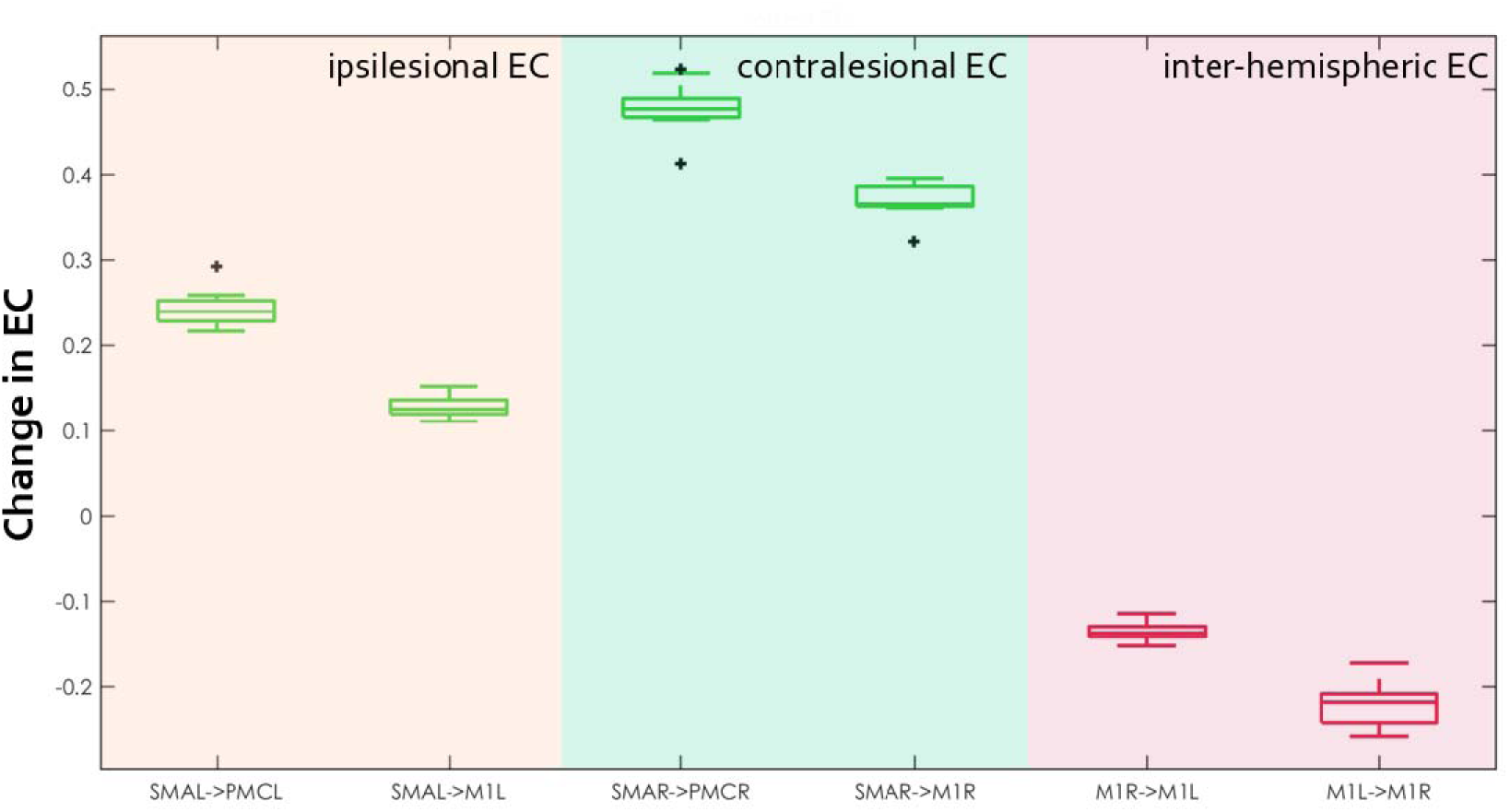
Changes in EC connectivity across patients for 6 representative connections. Boxplots indicate the median (central mark) and the 25th and 75th percentiles (bottom and top edges of the box) of the EC changes across 9 patients. Outliers are showed with a black cross. Green boxplots correspond to an increase in connectivity strength from b-s1 to b-s5; inversely red boxplots correspond to a decrease in EC after NF training.

## Discussion

We applied a DCM analysis to task-based fMRI data to describe the effect of NF training on bilateral motor networks in nine chronic stroke patients. The training protocol included five bimodal EEG-fMRI NF and nine EEG-NF sessions over five weeks aiming at reinforcing the activity of specific motor areas (ipsilesional SMA and M1). Results indicate that NF training induced a modulation of fMRI activation during the NF MI task towards ipsilesional motor cortex activation. We also found that the training elicited a reorganization of effective connectivity, with a general trend to reduced inter-hemispheric connectivity between premotor and primary motor cortices. These exploratory results suggest that the training of specific motor areas with NF can have an impact on the whole motor connectivity network.

### Methods

Different estimator of EC from fMRI data are available [60]. In this study, DCM was used for EC estimation because it was specifically developed for fMRI analysis. DCM has the advantage over approaches such as structural equation modeling or Granger causality that it uses a hemodynamic model to decompose the measured data into underlying neuronal signal and hemodynamic effects [49]. Moreover, DCM is particularly robust in dealing with deviations from the standard hemodynamic response, (e.g. due to pathology affecting blood flow parameters such as stroke) as the parameters in the hemodynamic model are estimated together with the parameters quantifying neuronal connectivity and individually for each ROI [49], [50]. In the definition of the *apriori* model of underlying connectivity patterns, we considered six regions of interest including both ipsilesional and contralesional premotor and motor areas. Other areas importantly involved in motor network (i.e. cerebellum, prefrontal areas) were excluded from the analysis as in DCM models including more than 8 ROIs, additional prior constraints are needed to reduce the number of parameters to estimate [61]. Moreover, DCM analysis does not result in erroneous estimations when regions that may have an influence on the model are disregarded, because information by brain regions not explicitly modelled is captured implicitly in the coupling parameters between two regions [62].

### fMRI activation and effective connectivity during a NF-MI task

Group level fMRI results yield activations of the areas typically involved during a MI task: SMA, PMC and posterior parietal cortex (PPC) [63]. These results are also in line with findings indicating that PPC is generally active when feedback is presented visually [64]. Activation patterns evolved from the first session to the last of the training. The first session involved more robustly SMA and the last session showed a larger recruitment of bilateral PMC and ipsilesional M1. SMA has been associated to complex MI tasks, involving a sequence of movements [63]. A higher SMA activation in the first NF session may indicate that at the beginning of the training patients were trying various MI tasks with more diversified motor planning, while at the end they were using the MI strategy learned in the course of the training. As also observed in our pilot study[25], giving general instructions without mentioning a precise MI task encouraged the patients to explore a range of different strategies before eventually focusing on a specific one.

Whether M1 is consistently activated during a MI task is debated. Functional MRI NF studies found non-conclusive results at group level [65], [66] and one recent work reported deactivation of M1 during MI training of the SMA and M1 [67]. A recent meta-analysis only reported consistent activation within the primary motor cortex during MI in a minority of studies (18%) [63]. The authors of this study suggest than only skilled MI performer may be able to activate M1 during a MI task, as found for instance by Sharma et al. [68], which could explain why M1 activation can be more consistently reported in single-subject analyses [69]. Our results indicated an activation of ipsilesional M1 in the last NF session suggesting on the one hand that patients “responded” to the NF reinforcement scheme targeting ipsilesional M1, and on the other that they became better at performing the MI task, which is to be expected after 15 motor NF training sessions.

Average connectivity results are in line with previous studies investigating EC patterns in stroke during MI or execution. They indicate a dense, bilateral connectivity pattern, with an inhibitory connection between contralateral M1 and ipsilesional M1. Connectivity in the contralesional hemisphere is commonly elicited in stroke patients during MI of the affected limb and there is evidence that the inhibitory influence from contralesional M1 to ipsilesional M1 increases in stroke patients with respect to healthy controls [13]. The role of contralesional M1 in stroke is debated as it has been shown to have both promoting and inhibitory influences on motor outcomes, depending on the severity and time from stroke (Gerloff et al., 2006; Grefkes et al., 2008b; Lotze et al., 2006; Murase et al., 2004; Nowak et al., 2008). For instance, a suppression of the contralesional M1 excitability has been shown to degrade upper limb control in severely impaired patients but to improve it for mildly impaired stroke patients [70].

### Effect of NF training on effective connectivity

Few recent studies have revealed the potential of modulating functional connectivity between the ipsilesional motor cortex and other cortical [34] or subcortical regions [33] for stroke rehabilitation. However, to the best of our knowledge, this is the first work that assesses fMRI connectivity changes resulting from the NF training of target motor areas in stroke. More generally, this work tackles the need to investigate how NF training of localized activity affects the related brain networks to guide more effective NF strategies based on physiologically relevant network targets and to gain a deeper insight into the underlying pathological processes [71].

DCM analysis revealed significant alterations in EC after NF training relative to baseline. Firstly, a general decrease in inter-hemispheric connections was observed, including a decrease in the inhibitory influence of the contralesional M1 on the ipsilesional PMC and M1 (not significant). This inhibitory influence through transcallosal connections reduces the motor output of the damaged hemisphere in patients with severe motor deficit [4], [72], and is part of a well-known “maladaptive” plasticity mechanism in stroke [73]. In a DCM study from Grefkes and colleagues on the effect of transcranial magnetic stimulation applied on contralesional M1, the negative coupling from contralesional M1 was absent after the treatment, and, more interestingly, this effect correlated with improvement in motor performance of the paretic hand [4]. Caution is however recommended when interpreting the changes in contralesional M1 connectivity because, even if its alterations in stroke are well-documented, their influence on motor recovery is debated and insufficiently understood [1].

We observed an increase in feedforward connectivity strength between ipsilesional SMA and PMC as a result of the NF training. Using a similar BMA approach, Bajaj and colleagues compared the connectivity before and after mental practice and physical therapy intervention [14]. In line with our findings, they reported an increase in EC between SMA and PMC in the affected hemisphere during MI task and this increase was significantly correlated with the motor score improvement after treatment. Similarly, Sharma and colleagues reported that coupling between PMC and SMA is diminished in stroke patients with respect to healthy controls and that as motor function improved, the coupling between these areas increased [74].

The general reduction of connectivity from ipsilesional to contralesional motor areas (see also Figure S4 in Supplementary Material) suggests a pattern closer to a unilateral motor imagery network in healthy subjects. The inhibitory influence from M1 to the contralateral motor areas is a well described mechanism of unilateral hand movements [75] and an increase in inhibitory coupling from ipsilesional to contralesional M1 may indicate a more lateralized MI of the affected limb. Moreover, an increase in the inhibitory coupling from M1 to SMA and, inversely, a positive coupling from SMA to M1 corresponds to the feed-forward connectivity model estimated with DCM in healthy controls in [76]. More generally, the observed trend of reduction in connectivity can be also interpreted as an effect of learning. Average EC patterns show a fully connected bilateral network during MI (Figure 3A): a decrease in the strength of EC suggest that the same MI task could be performed with a less dense network at the end of the NF training. A similar trend was observed in connectivity patterns during language prosody NF training where connectivity was reduced and more localized at the end of the training [77].

Finally, the decrease in self-inhibitory connections in ipsilesional M1 and bilateral SMA, indicate that these regions were more activated at the end of the NF training. This is in line with the results of the fMRI activation analysis and, more interestingly, with the NF reinforcement scheme that rewarded upregulation of ipsilesional SMA and M1.

### Future work

The population of this study includes stroke patients with a relatively broad range of stroke latency and lesions localization. Individual motor and structural impairment differences following stroke might induce variability to the estimation of EC with DCM. Unfortunately, this is the case of the majority of fMRI-NF studies, whose sample size is strongly limited by the cost and burden of MRI imaging and also true in this case, considered the technical challenge of bimodal EEG-fMRI NF and the fact that patients underwent a long training program of 14 NF sessions. In future, a larger sample of patients could be considered to refine our findings of NF training effect on connectivity networks and extend our observations.

Building on these first results, in future work we will also assess if the connectivity changes elicited by NF training resulted in improved motor performance of the affected limb, taking clinical outcomes into account. Our current analysis could then be extended to the whole sample of our randomized controlled study, and network reorganization patterns could then be compared between the interventional and control group.

Overall, we believe that this work represents a valuable and novel analysis of the large-scale effects of localized NF training on fMRI connectivity, and, to the best of our knowledge, the first in stroke. It underlines the importance of investigating also connectivity patterns rather than focusing on regional activation when assessing NF training outcomes.

## Conclusions

Using a DCM approach, in this work we investigate the effect of multi-session EEG-fMRI NF training on connectivity patterns in chronic stroke patients. The DCM model consisted in a bilateral motor network including premotor, supplementary and primary motor areas and a Bayesian Model Average approach was used to assess significant changes in connectivity following the NF training. Results show that upregulation of ipsilesional M1 and SMA modulates fMRI activity and effective connectivity in both hemispheres and generally reduces inter-hemispheric connectivity strength. Our results suggest that the upregulation of target motor areas by means of NF can have a larger scale impact on connectivity and a potential to mitigate maladaptive networks patterns.

## Data Availability

N.A.

## Acknowledgements

MRI data acquisition was supported by the Neurinfo MRI research facility from the University of Rennes I. Neurinfo is granted by the the European Union (FEDER), the French State, the Brittany Council, Rennes Metropole, Inria, Inserm and the University Hospital of Rennes. We thank Gabriela Vargas for her useful suggestions for the DCM analysis. The project was supported by the National Research Agency in the “Investing for 540 the Future” program under reference ANR-10-LABX-07-0, and by the “Fondation pour la Recherche Médicale” under the convention #DIC20161236427.

## Author contributions

G.L. wrote the manuscript, A.V. and Q.D. contributed to methods section writing. G.L., I.B., S.B., C.B. and A.L. designed the studies. I.B., S.B. and E.L. contributed to patient enrollment and screening. A.V., G.L., J.C. and Q.D. performed the data analysis. G.L., E.L. and M.F. collected the data. G.L. and E.B. contributed to the design of the experimental platform. All authors reviewed the manuscript.

